# When two communication differences intersect: Comparing inpatient rehabilitation care and outcomes for people with post-stroke aphasia who do and do not require an interpreter

**DOI:** 10.1101/2024.05.20.24307645

**Authors:** Kathleen Mellahn, Monique Kilkenny, Samantha Siyambalapitiya, Ali Lakhani, Catherine Burns, Dominique A. Cadilhac, Miranda L. Rose

## Abstract

**Background:** Communicative ability after stroke influences patient outcomes. Limited research has explored the impact of aphasia when it intersects with cultural or linguistic differences on receiving stroke care and patient outcomes. We investigated associations between requiring an interpreter and the provision of evidence-based stroke care and outcomes for people with aphasia in the inpatient rehabilitation setting.

**Methods:** Patient-level data from people with aphasia were aggregated from the Australian Stroke Foundation National Stroke Audit - Rehabilitation Services (2016-2020). Multivariable regression models compared adherence to processes of care (e.g. home assessment complete, type of aphasia management) and in-hospital outcomes (e.g. length of stay, discharge destination) by requirement of an interpreter. Outcome models were adjusted for sex, stroke type, hospital size, year, and stroke severity factors.

**Results:** Among 3160 people with aphasia (median age 76, 56% male), 208 (7%) required an interpreter (median age 77, 52% male). The interpreter group had more severe disability on admission, reflected by reduced cognitive (6% vs 12%, p<0.0000) and motor FIM scores (6% vs 12%, p<0.009). The interpreter group were less likely to have phonological and semantic interventions for their aphasia (OR 0.56, 95% CI 0.40, 0.78) compared to people not requiring an interpreter. They more often had a carer (OR 2.03, 95% CI 1.41, 2.96) and were less likely to have a home assessment prior to discharge (OR 0.34, 95% CI 0.12, 0.95) despite increased likelihood of discharging home with supports (OR 1.49, 95% CI 1.08, 2.05). The interpreter group had longer lengths of stay (median 31 vs 26 days, p=0.005).

**Conclusion:** Some processes of care and outcomes differed in inpatient rehabilitation for people with post-stroke aphasia who required an interpreter compared with those who did not. Equitable access to therapy is imperative and greater support for cultural/linguistic minorities during rehabilitation is indicated.

## INTRODUCTION

Inefficiency and breakdown in communication can impact healthcare delivery regardless of the medical presentation or clinical setting. ^1, 2^ This is particularly significant when there is discordance in communication between the service provider and the patient. Two examples of these communication differences are communication disability and having a culturally and linguistically diverse (CALD) background.^3–6^

Aphasia is a language disorder and communication disability impacting approximately 30% of people who have a stroke. ^7, 8^ People with post-stroke aphasia have greater risks of adverse events and longer lengths of stay in hospital than their peers without aphasia. ^3, 6, 9^

Minority cultural or linguistic groups may also experience challenges to care, especially if they have limited English proficiency when treated in an English-speaking country. ^10–13^ Globalization and high levels of migration have led to increasingly multicultural populations, especially in Western countries such as Australia, the United Kingdom, Canada and the United States of America.^14–17^ Regarding stroke care for CALD populations, Rezania et al. noted disparities in thrombolysis access, discharge destinations, and length of stay. ^4^ After hospital discharge, two studies found that post-stroke independence was less likely for those from CALD backgrounds, particularly when an interpreter was needed.^10, 18^

Until recently exploration of the compounded disadvantage experienced by people who have aphasia and are from a CALD population has been limited. Our previous research found differences in receiving evidence-based acute stroke care for people with aphasia who required an interpreter compared to those not requiring an interpreter.^19^ It remains unclear if differences also exist during inpatient rehabilitation.

Investigating neurorehabilitation differences for post-stroke aphasia is crucial, given the importance of task and stimuli salience, and patient motivation on experience-dependent neuroplasticity. A double-layered communication barrier (CALD; aphasia) may significantly affect the therapeutic relationship and rehabilitation quality, potentially impeding community re-integration. ^20, 21^

The aim of this study was to investigate associations between requiring an interpreter and the provision of evidence-based stroke care and outcomes for people with aphasia in Australian inpatient rehabilitation settings.

## METHODS

This is a cross-sectional observational study using pooled national stroke audit data that conforms to the STrengthening the Reporting of OBservational studies in Epidemiology (STROBE) Guidelines for cross-sectional studies.^22^ We used data collected in the biennial Stroke Foundation National Stroke Audit for Rehabilitation services conducted in 2016, 2018 and 2020.^23–25^ The Audit Program comprises two components: an organizational survey completed by a trained staff member familiar with the stroke rehabilitation service; and a clinical audit questionnaire of up to 40 consecutive stroke rehabilitation admissions.^26^ The organizational survey collects data about the structure of the stroke rehabilitation workforce and resources, and the clinical audit questionnaire extracts patient-level data concerning hospital outcomes and evidence-based clinical care – referred to as processes of care. Thorough details of the Audit Program methodology have been published previously. ^26^ The dependent variables collected were grouped into four categories: admission characteristics, discipline specific processes of care, discharge planning processes of care and in-hospital outcomes: the specific details of these can be found in Supplemental Table 1. Admission characteristics included demographic information such as age and sex; pre-transfer location; rehabilitation ward type and hospital size; stroke type; stroke severity indicators on admission; and level of independence (modified Rankin Scale [mRS], Cognitive and Motor components of Functional independence Measure [FIM]). We gathered data on First Nations identity, although the CALD label typically excludes First Peoples. They were included here due to the criterion of requiring an interpreter. Discipline specific processes of care included assessment by allied health disciplines, and the type of management for mobility, activities of daily living (ADLs), neglect, upper limb, aphasia, and malnutrition. Discharge planning processes of care consisted of development of a discharge care plan; preparation for discharge such as carer training and home assessment; discussion about returning to driving and work; and communication and peer support on discharge such as being provided a hospital contact. In-hospital outcomes comprised complications during admission, discharge destination, independence on disability measures, and length of stay.

Consistent with our research aims, we exclusively extracted data for the patients recorded as having aphasia in the clinical audit. CALD status was identified by whether the patient required an interpreter, as country of birth and ethnicity were not collected in the Audit Program. Although this is not an ideal measure it has been used in other health research^27^. Aggregated data from all hospitals participating in any of the audit cycles were included.

### Data Analysis

Demographic and admission clinical information, processes of care and in-hospital outcomes for people with post-stroke aphasia were compared for patients who did and did not require an interpreter as detailed below. Where questions related to admission characteristics or in-hospital outcomes, ‘not documented’ or ‘unknown’ responses were both assumed to be negative and included in the denominator for the analysis. Records with missing responses for processes of care were considered incomplete and excluded from analysis. Data were analyzed using STATA SE 18.0 (StataCorp, Texas).

Descriptive statistics were used to characterize all variables of interest by interpreter status. Between group differences were compared using Kruskal-Wallis and chi square tests as appropriate. All statistical tests were two-sided, with level of significance at p<0.05. Demographic and admission characteristics that demonstrated a statistically significant difference, were analyzed for collinearity with factors known to influence outcomes (age, sex, pre-morbid independence [mRS 0-2] stroke type, and stroke severity [arm and mobility impairment]). Multivariable logistic regression models were used to analyze non-collinear variables for processes of care and in hospital outcomes. If collinearity was demonstrated the variable with greatest evidence for impacting stroke outcomes based on current literature would have been chosen. Audit year was included in the model as a sensitivity analysis.

The final regression model utilized in this study adjusted for age, sex, pre-morbid independence, stroke type, audit year, arm and mobility impairments, hospital size and independence on admission on each of the Cognitive and Motor components of the FIM.

When assessing between group differences for processes of care and in-hospital outcomes that had a p value of less than 0.1 on either chi square or Kruskal Wallis tests underwent multivariable, multilevel logistic regression with level defined as hospital to determine differences by interpreter status.

Random effects logistic regression was used for binary outcomes or processes of care (e.g., seen by a Physiotherapist, independence on discharge), including the outcome or process of care as a dependent variable in separate models, with level being hospital. Median regression models were used for length of stay, with clustering to account for hospital differences. Results of multivariable models were reported as adjusted odds ratios (aORs) or coefficients, and 95% confidence intervals (CIs).

## RESULTS

### Demographic and clinical characteristics

Overall, 10007 rehabilitation admissions were audited from 133 hospitals across the three audit cycles, whereby 3160 (32%) patients were documented to have aphasia. In the aphasic cohort, 208 (7%) required an interpreter.

On admission, the interpreter group were admitted to rehabilitation later after stroke onset (median 10 days vs 9 days, p=0.0038), were more likely to be admitted to a mixed rehabilitation unit (i.e. not strictly neurorehabilitation) and be admitted to hospitals that treated more than 80 stroke patients a year (Table 1). Although there were no differences in stroke type, the interpreter group were more likely to have a mobility impairment and were less likely to be independent on all three disability outcome measures (mRS, FIM Cognition and FIM Motor). The interpreter group were less likely to be employed at time of admission (10% vs 19% no interpreter) and more likely to have a carer as a support person (68% vs 48%).

**Table 1.**
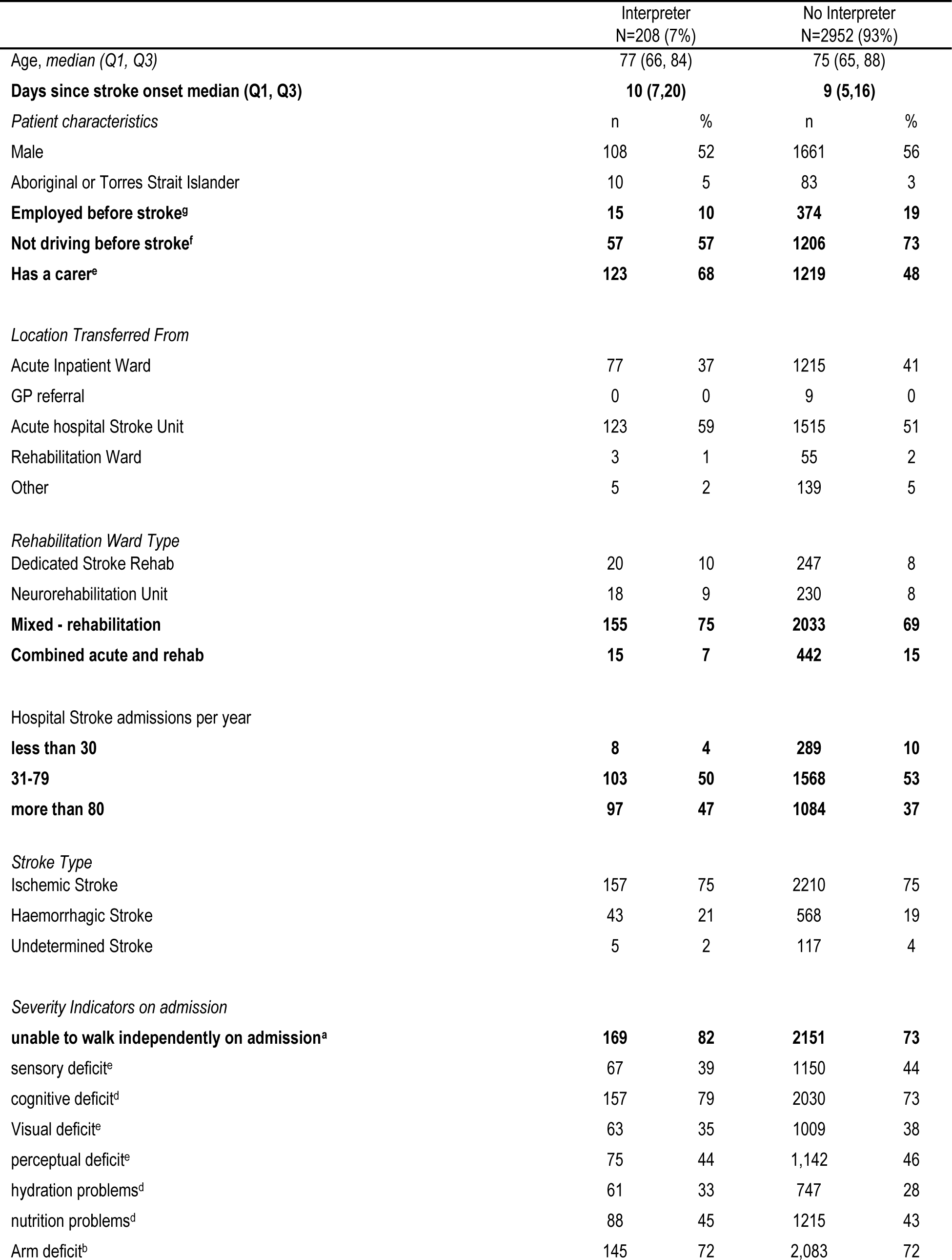
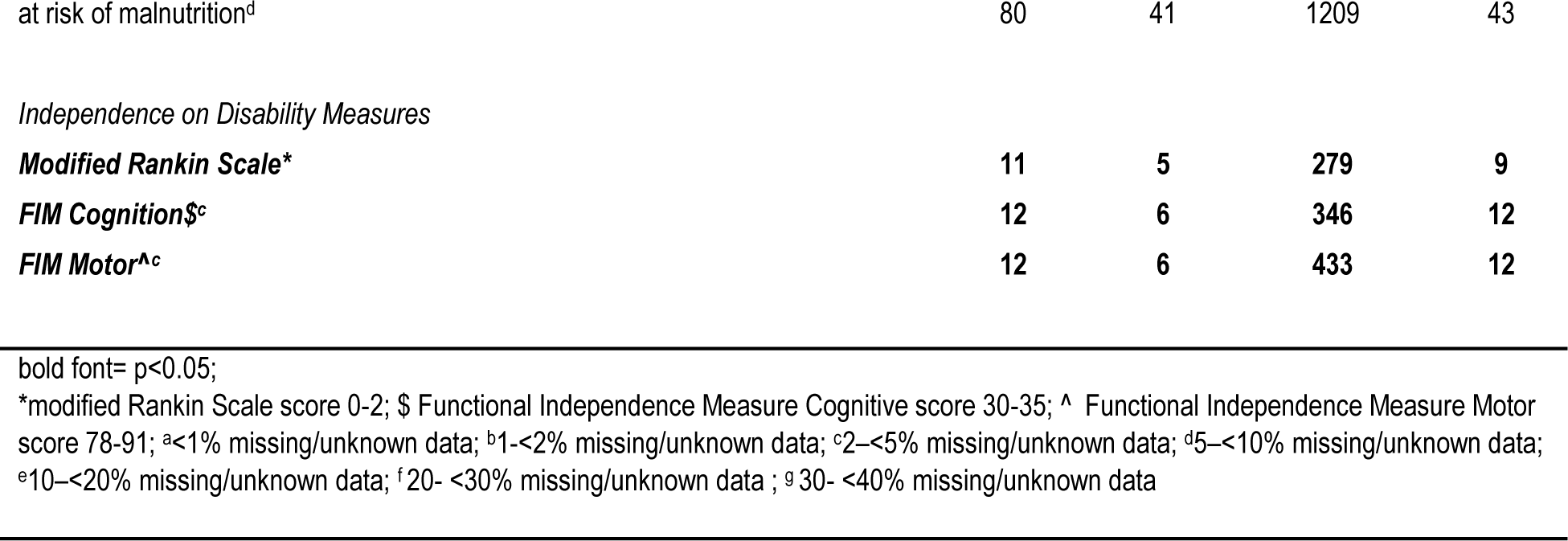
Demographics and characteristics of patients with aphasia by interpreter status on admission to inpatient rehabilitation ward.

Although univariable analysis found differences in proportions between groups across processes of care and in-hospital outcomes that often favored the group not requiring an interpreter, few aspects of care or outcomes remained significantly different in the multivariable analyses (Tables 1-4).

### Processes of Care

After multi-variable analysis, post-stroke impairment management was seen to be different, for neglect and aphasia management (Table 2). We found that the interpreter group were less likely to have visual scanning (aOR 0.50, 95% CI 0.28, 0.88) utilized in the management of their neglect. Those requiring an interpreter were also less likely to have phonological or semantic interventions (aOR 0.57, 95% CI 0.40, 0.80); constraint induced language therapy (aOR 0.33, 95% CI 0.15, 0.72) or group therapy (aOR 0.47, 95% CI 0.30, 0.73) than those who did not require an interpreter.

**Table 2.**
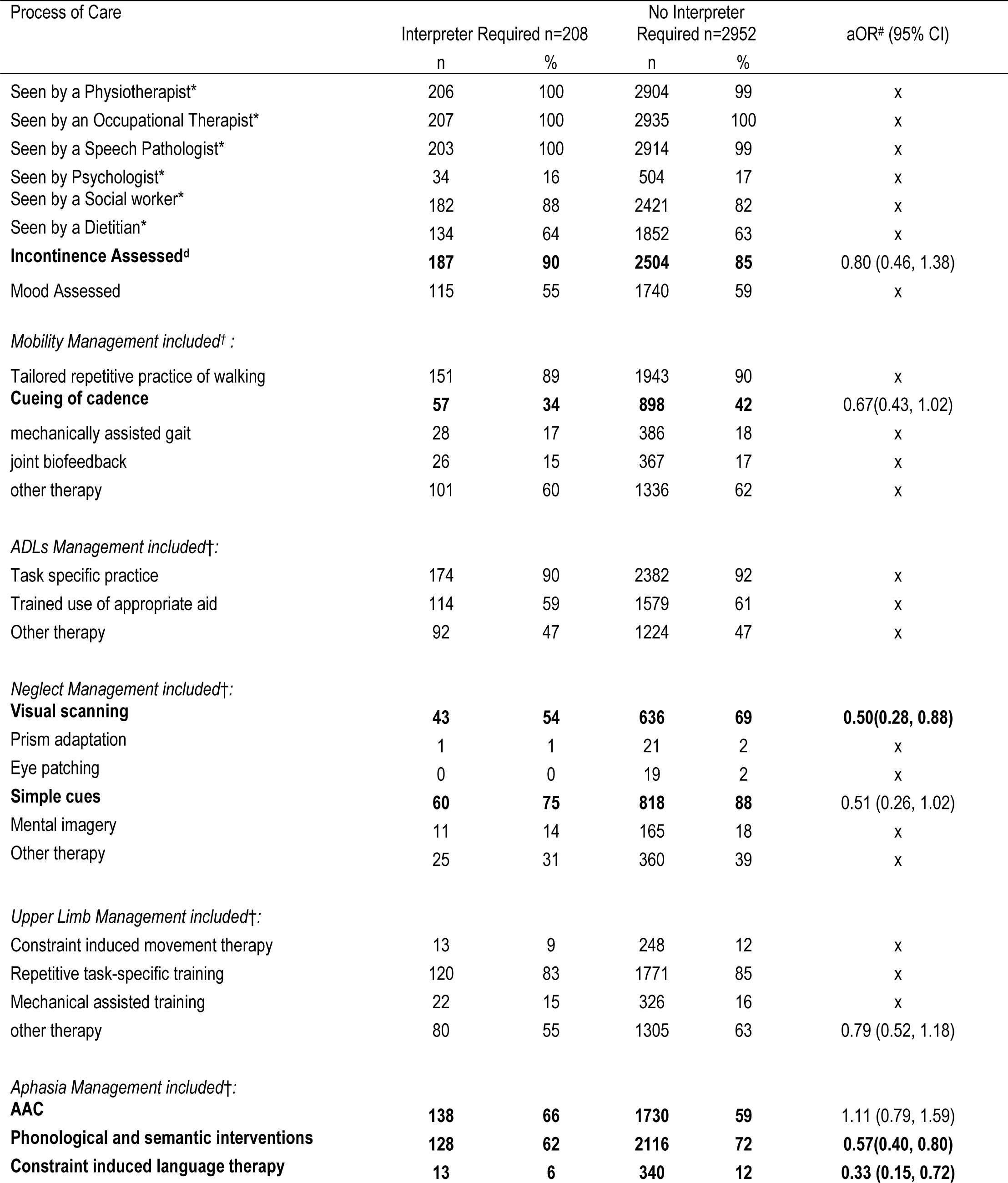
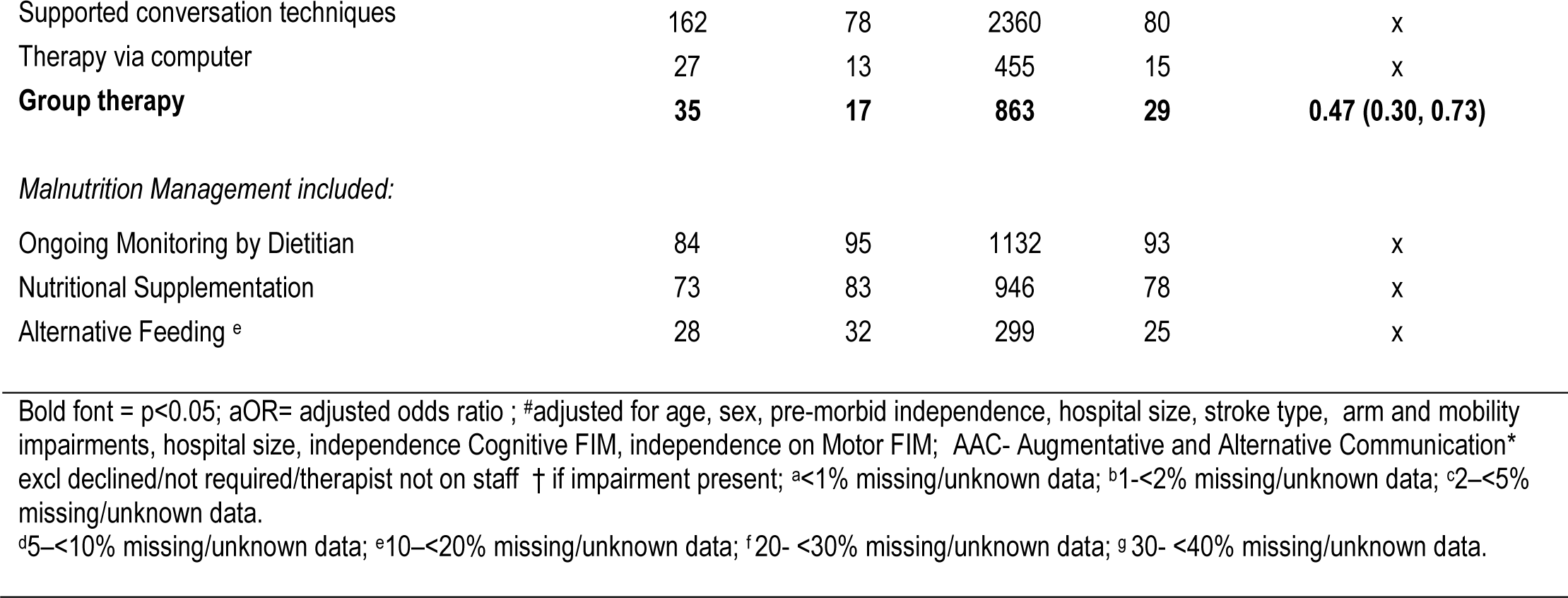
Adherence to Discipline specific processes of care by interpreter status.

| Process of Care | Interpreter Required n=208 |  | No Interpreter Required n=2952 |  | aOR<sup>#</sup> (95% CI) |
| --- | --- | --- | --- | --- | --- |
|  | n | % | n | % |  |
| Seen by a Physiotherapist* | 206 | 100 | 2904 | 99 | x |
| Seen by an Occupational Therapist* | 207 | 100 | 2935 | 100 | x |
| Seen by a Speech Pathologist* | 203 | 100 | 2914 | 99 | x |
| Seen by Psychologist* | 34 | 16 | 504 | 17 | x |
| Seen by a Social worker* | 182 | 88 | 2421 | 82 | x |
| Seen by a Dietitian* | 134 | 64 | 1852 | 63 | x |
| **Incontinence Assessed<sup>d</sup>** | **187** | **90** | **2504** | **85** | 0.80 (0.46, 1.38) |
| Mood Assessed | 115 | 55 | 1740 | 59 | x |
| *Mobility Management included<sup>†</sup>:* |  |  |  |  |  |
| Tailored repetitive practice of walking | 151 | 89 | 1943 | 90 | x |
| **Cueing of cadence** | **57** | **34** | **898** | **42** | 0.67(0.43, 1.02) |
| mechanically assisted gait | 28 | 17 | 386 | 18 | x |
| joint biofeedback | 26 | 15 | 367 | 17 | x |
| other therapy | 101 | 60 | 1336 | 62 | x |
| *ADLs Management included<sup>†</sup>:* |  |  |  |  |  |
| Task specific practice | 174 | 90 | 2382 | 92 | x |
| Trained use of appropriate aid | 114 | 59 | 1579 | 61 | x |
| Other therapy | 92 | 47 | 1224 | 47 | x |
| *Neglect Management included<sup>†</sup>:* |  |  |  |  |  |
| **Visual scanning** | **43** | **54** | **636** | **69** | **0.50(0.28, 0.88)** |
| Prism adaptation | 1 | 1 | 21 | 2 | x |
| Eye patching | 0 | 0 | 19 | 2 | x |
| **Simple cues** | **60** | **75** | **818** | **88** | 0.51 (0.26, 1.02) |
| Mental imagery | 11 | 14 | 165 | 18 | x |
| Other therapy | 25 | 31 | 360 | 39 | x |
| *Upper Limb Management included<sup>†</sup>:* |  |  |  |  |  |
| Constraint induced movement therapy | 13 | 9 | 248 | 12 | x |
| Repetitive task-specific training | 120 | 83 | 1771 | 85 | x |
| Mechanical assisted training | 22 | 15 | 326 | 16 | x |
| other therapy | 80 | 55 | 1305 | 63 | 0.79 (0.52, 1.18) |
| *Aphasia Management included<sup>†</sup>:* |  |  |  |  |  |
| **AAC** | **138** | **66** | **1730** | **59** | 1.11 (0.79, 1.59) |
| **Phonological and semantic interventions** | **128** | **62** | **2116** | **72** | **0.57(0.40, 0.80)** |
| **Constraint induced language therapy** | **13** | **6** | **340** | **12** | **0.33 (0.15, 0.72)** |
| Supported conversation techniques | 162 | 78 | 2360 | 80 | x |
| Therapy via computer | 27 | 13 | 455 | 15 | x |
| **Group therapy** | **35** | **17** | **863** | **29** | **0.47 (0.30, 0.73)** |
| *Malnutrition Management included:* |  |  |  |  |  |
| Ongoing Monitoring by Dietitian | 84 | 95 | 1132 | 93 | x |
| Nutritional Supplementation | 73 | 83 | 946 | 78 | x |
| Alternative Feeding <sup>e</sup> | 28 | 32 | 299 | 25 | x |

Bold font = p<0.05; aOR= adjusted odds ratio ; #adjusted for age, sex, pre-morbid independence, hospital size, stroke type, arm and mobility impairments, hospital size, independence Cognitive FIM, independence on Motor FIM; AAC- Augmentative and Alternative Communication\* excl declined/not required/therapist not on staff † if impairment present; <sup>a</sup><1% missing/unknown data; <sup>b</sup>1-<2% missing/unknown data; <sup>c</sup>2-<5% missing/unknown data.
<sup>d</sup>5-<10% missing/unknown data; <sup>e</sup>10-<20% missing/unknown data; <sup>f</sup>20- <30% missing/unknown data; <sup>g</sup>30- <40% missing/unknown data.

Several between group differences were found for discharge planning processes of care (Table 3). Compared to those not requiring an interpreter, the interpreter group were less likely to have a home assessment completed (aOR 0.34, 95% CI 0.12, 0.95) or be asked if they wanted to return to driving (aOR 0.33, 95% CI 0 .20, 0.55). Requiring an interpreter was associated with a decreased likelihood of being offered information about sexuality after stroke (aOR 0.45, 95% CI 0.24, 0.83) or self-management programs (aOR 0.41, 95% CI 0.18, 0.91).

**Table 3.**
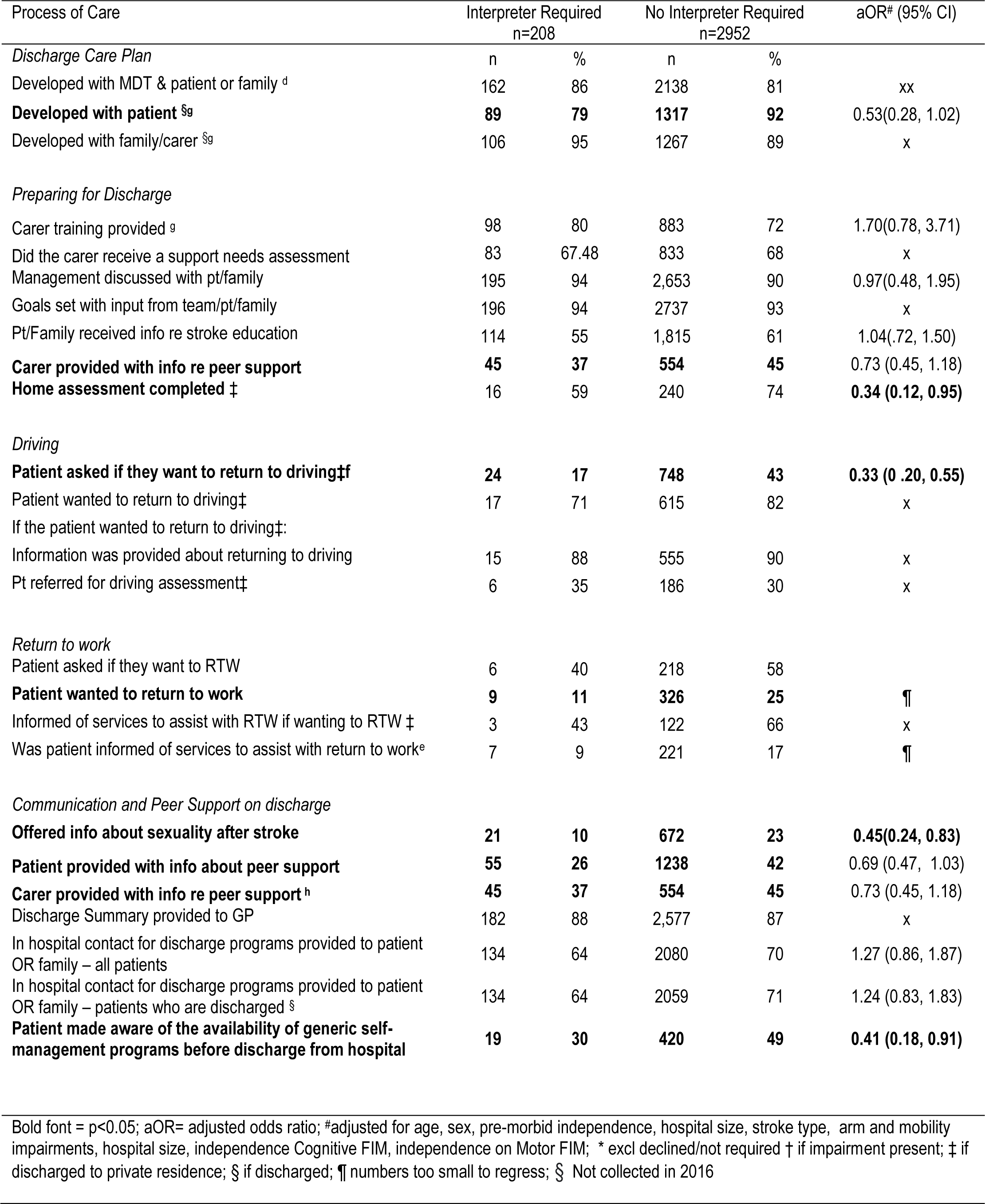
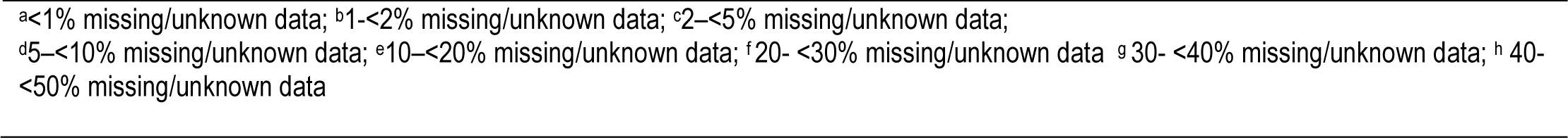
Adherence to discharge planning processes of care by interpreter status.

**Table 4.** In-hospital outcomes by interpreter status for people with aphasia.

| Outcome | Interpreter Required<br>n=208 |  | No Interpreter Required n=2952 |  | aOR# (95% CI) |
| --- | --- | --- | --- | --- | --- |
| *Complications during admission* | n | % | n | % |  |
| Deaths | 0 | 0 | 43 | 1 | x |
| aspiration pneumonia | 14 | 7 | 175 | 6 | x |
| DVT | 2 | 1 | 44 | 1 | x |
| Fall | 41 | 20 | 461 | 16 | x |
| Fever | 24 | 12 | 279 | 9 | x |
| pressure sore | 12 | 6 | 143 | 5 | x |
| shoulder subluxation | 14 | 7 | 172 | 6 | x |
| shoulder pain | 29 | 14 | 395 | 12.38 | x |
| UTI<sup>a</sup> | 36 | 17 | 474 | 16.09 | x |
| Contracture | 6 | 3 | 59 | 2 | x |
| Malnutrition | 21 | 10 | 323 | 10.94 | x |
| new AF<sup>g</sup> | 5 | 4 | 81 | 4.05 | x |
| *Discharge Destination* |  |  |  |  |  |
| Other acute hospital | 13 | 6 | 296 | 10 | 0.72 (0.40, 1.32) |
| New residential aged care service | 36 | 17 | 418 | 14 | x |
| Statistical discharge (type change) | 8 | 4 | 102 | 4 | x |
| Other | 6 | 3 | 59 | 2 | x |
| Left against medical advice | 2 | 1 | 34 | 1 | x |
| **Usual residence with supports** | **118** | **57** | **1419** | **49** | **1.48(1.08, 2.04)** |
| Usual residence with NO supports | 15 | 7 | 279 | 10 | x |
| Inpatient rehabilitation | 3 | 1 | 98 | 3 | x |
| **Transitional care services** | **7** | **3** | **204** | **7** | **0.40 (0.17, 0.94)** |
| *Independence on Disability Measures* |  |  |  |  |  |
| **Modified Rankin Scale<sup>\*A</sup>** | **77** | **37** | **1399** | **47** | 0.80 (0.52, 1.25) |
| **FIM Cognition<sup>\$</sup>** | **25** | **13** | **740** | **27** | **0.48 (0.28, 0.82)** |
| **FIM Motor<sup>^d</sup>** | **62** | **32** | **1216** | **44** | 0.81 (0.55, 1.18) |
|  | Interpreter Required<br>n=208 |  | No Interpreter Required n=2952 |  |  |
|  | median | Q1,Q3 | median | Q1,Q3 |  |
| *Change in FIM* |  |  |  |  |  |
| *FIM Motor<sup>^</sup>* | 19 | 7,31 | 17 | 6,30 | x |
| *FIM Cognition<sup>\$</sup>* | 3 | 0, 7 | 3 | 0, 7 | x |
| *Length of Stay* |  |  |  |  | Coefficient (95% CI) |
| **Including Deaths** | **31** | **18, 46** | **26** | **14,42** | **4.5 (1.25, 7.75)** |
| **Excluding Deaths** | **31** | **18, 46** | **26** | **14,42** | **3 (-0.23, 6.23)** |

Bold font = p<0.05; aOR= adjusted odds ratio ##adjusted for age, sex, pre-morbid independence, hospital size, stroke type, arm and mobility impairments, hospital size, independence Cognitive FIM, independence on Motor FIM ; ¶ numbers too small to regress. ; \* defined as modified Rankin Scale score 0-2; \$ defined as Functional Independence Measure Cognitive score 30-35; ^ defined as Functional Independence Measure Motor score 78-91; <sup>a</sup><1% missing/unknown data; <sup>c</sup>2-<5% missing/unknown data; <sup>d</sup>5-<10% missing/unknown data; ; <sup>g</sup> 30- <40% missing/unknown data;

### In-hospital outcomes

Compared to those not requiring an interpreter, the interpreter group were more likely to discharge home with supports (aOR 1.48, 95% CI 1.08, 2.04) and less likely to discharge to a transitional care service (aOR 0.40, 95% CI 0.17, 0.94) (Table 4). In terms of disability outcome measures, patients requiring an interpreter were less likely to be independent on all measures (mRS, FIM Cognition and FIM Motor). However, following multivariable analyses, only independence on FIM Cognition remained statistically significantly different between groups (aOR 0.48, 95% CI 0.28, 0.82).

Finally, requiring an interpreter was associated with a longer median rehabilitation stay (31 days vs. 26 days). After median regression, LOS difference remained significant: the interpreter group had a coefficient of 4.5 (p=0.007), indicating a 4.5-day longer stay than peers without interpreter needs.

## Discussion

To our knowledge, quantitative analyses comparing the inpatient rehabilitation care of people with aphasia from CALD backgrounds to their non-CALD peers have not been reported. We found that people with aphasia who required an interpreter had different clinical profiles on admission to rehabilitation. First, the interpreter group were more likely to be treated in larger hospitals in metropolitan regions which could stem from historical migration trends favoring major Australian cities. ^28^ Our data showed that people with aphasia with interpreter needs arrived at rehabilitation units later post-stroke, were more likely to have a carer, less likely to be independent on disability measures and were more likely to be unable to walk. Each of these findings align with acute research, which showed that requiring an interpreter was associated with a 2-day longer length of stay and a decreased likelihood of independence upon discharge.^4, 19^

Our data showed some differences in evidence-based stroke rehabilitation provision that were not attributable to differences in admission disability severity, or other confounding factors. Requiring an interpreter was associated with reduced access to impairment-based and group aphasia therapies which may be explained by the difficulty of providing communication therapy when there is a language discordance between clinician and patient.^29^ Factors contributing to this difficulty include limited culturally and linguistically appropriate therapy resources, and logistical challenges in coordinating interpreters. Differences in care extended beyond speech pathology with the interpreter group being half as likely to receive visual scanning for the management of neglect, typically managed by Occupational Therapists.^30, 31^

Differences were also evident in discharge planning. The interpreter group were two thirds less likely to have a home assessment completed or to be asked if they wanted to return to driving. Similarly, they were less than half as likely to be offered information about sexuality after stroke or to be made aware of general self-management programs. Lack of appropriate translated resources to provide this information and assessment could hinder these processes.^32^

Finally, we observed significant outcome differences between groups. Requiring an interpreter was associated with a 4.5 day longer length of stay. Despite protracted admissions, the interpreter group were less independent on discharge on all three disability outcomes. For modified Rankin Scale and FIM Motor the statistical significance of this difference was explained by disability severity on admission. However, for FIM Cognition, admission disability severity could not account for differences in cognitive independence on discharge. Interpreter status influenced discharge destination, with these patients more likely to transition home with support and less likely to be discharged to a transitional care program, notwithstanding reduced rates of home assessment. This may be linked to the higher rates of available carers for the interpreter group meaning discharge home was more likely to be perceived as a viable option.

### Strengths and Limitations

Previous work has detailed the strengths and limitations of the National Stroke Audit Program data. ^19^ Strengths include its quality and representative nature; assured by a large dataset and reliable collection processes. This involves trained data abstractors, a data dictionary, and a web-based entry tool with built-in logic checks to minimize errors.

Despite these advantages, the imbalance in group sizes and retrospective, cross-sectional nature of the data may limit our findings’ power. Given this data is from an Australian, largely public hospital setting, caution is needed when drawing comparisons or generalizing to stroke rehabilitation more broadly.

The main limitation of this data set is the limited options for identifying CALD status.^19^ Additional demographic details such as country of birth, languages spoken, and parental country of birth should be collected to better differentiate CALD from non-CALD groups, allowing for a more nuanced analysis of the influence of culture and ethnicity on care. ^33^ Our research suggests that the interpreter group represent a particularly marginalized population, as they are potentially less likely to be bilingual. This may explain the small proportion of patients (7%) documented to require an interpreter when census data suggests the number of people who speak languages other than English should be closer to 20%.^17^

While a large range of information was extracted, gaps in our understanding of the clinical picture persist due to the limitations on data collected in the Audit Program. Specifically, we lack data on the frequency and quality of interventions, as well as whether interpreters were utilized during admission. Consequently, we cannot ascertain how the use of interpreters may influence differences in care and outcomes.

This work expands on our acute research and demonstrates that disparities in the clinical profile, provision of evidence-based care, and clinical outcomes extend into rehabilitation. ^19^ We have identified areas of compromised care quality which presents an opportunity for targeted improvement of stroke services; however, this requires further research that explores the decision-making underpinning clinicians’ management of CALD people with aphasia. Furthermore, more robust clinical data on interpreter use would enhance understanding and opportunities for practice change. Research is currently underway to address these knowledge gaps.

## Conclusions

Requiring an interpreter was found to critically influence several differences in the provision of inpatient rehabilitation and in-hospital outcomes for people with post-stroke aphasia. This highlights the necessity for enhanced support for CALD people to ensure equitable access to therapy. Further research is required to explore the system-level factors driving these differences to make meaningful practice changes and prevent service inequity.

## Data Availability

All data referred to in the manuscript is accessible stored and accessible by contacring The Stroke Foundation (Australia). Reports on the audit cycles and their respective data a freely available on The Stroke Foundation Website

https://informme.org.au/stroke-data/rehabilitation-audits

## Acknowledgements

We acknowledge the hospitals and clinicians participating in the National Stroke Audit Program.

## Ethical approval

Ethics approval for data used in this project was granted through the Human Research Ethics Committee from Monash University (Project ID 35037).

---

**Supplemental Table S1:** Variables collected for analysis.

| Admission Characteristics | Discipline Specific Processes of Care | Discharge Planning Processes of Care | In-Hospital Outcomes |
| --- | --- | --- | --- |
| Age | Seen by a Physiotherapist | *Discharge Care Plan*<br>Developed with MDT & patient or family<br>Developed with patient<br>Developed with family/carer | *Complications during admission:* |
| Sex | Seen by an Occupational Therapist |  | Death |
| Days since stroke onset on admission | Seen by a Speech Pathologist | *Preparing for Discharge:* | aspiration pneumonia |
|  | Seen by Psychologist | Carer training provided | DVT |
| Aboriginal or Torres Strait Islander | Seen by a Social worker | Did the carer receive a support needs assessment | Fall |
| Employed before stroke | Seen by a Dietitian | Management discussed with pt/family | Fever |
| Not driving before stroke | Incontinence Assessed | Goals set with input from team/pt/family | pressure sore |
| Has a carer | Mood Assessed | Pt/Family received info re stroke education | shoulder subluxation |
|  |  | Carer provided with info re peer support | shoulder pain |
| *Location Transferred From*<br>Acute Inpatient Ward<br>GP referral<br>Acute hospital Stroke Unit<br>Rehabilitation Ward<br>Other | *Mobility Management included†:*<br>Tailored repetitive practice of walking<br>Cueing of cadence<br>mechanically assisted gait<br>joint biofeedback<br>other therapy | Home assessment completed | UTI |
| *Rehabilitation Ward Type*<br>Rehabilitation Ward Type<br>Dedicated Stroke Rehab<br>Neurorehabilitation Unit<br>Mixed - rehabilitation<br>Combined acute and rehab | *ADLs Management included†:*<br>Task specific practice<br>Trained use of appropriate aid<br>Other therapy | *Driving:* | Contracture |
| Hospital Stroke admissions per year<br>less than 30<br>31-79<br>more than 80 | *Neglect Management included†:*<br>Visual scanning<br>Prism adaptation<br>Eye patching<br>Simple cues | Asked if they want to return to driving | Malnutrition |
|  | • Mental imagery<br>• Other therapy |  |  |
| Stroke Type<br>• Ischemic Stroke<br>• Haemorrhagic Stroke<br>• Undetermined Stroke | Upper Limb Management included†:<br>• Constraint induced movement therapy<br>• Repetitive task-specific training<br>• Mechanical assisted training<br>• other therapy | Patient wanted to return to driving | new atrial fibrillation |
| Severity Indicators on admission | Aphasia Management included†:<br>• AAC<br>• Phonological and semantic interventions<br>• Constraint induced language therapy<br>• Supported conversation techniques<br>• Therapy via computer<br>• Group therapy | If the patient wanted to return to driving: |  |
| unable to walk independently on admission | Malnutrition Management included:<br>• Ongoing Monitoring by Dietitian<br>• Nutritional Supplementation<br>• Alternative Feeding | Information was provided about returning to driving | Discharge Destination:<br>• Other acute hospital<br>• New residential aged care service<br>• Statistical discharge (type change)<br>• Other<br>• Left against medical advice<br>• Usual residence with supports<br>• Usual residence with NO supports<br>• Inpatient rehabilitation<br>• Transitional care services |
| sensory deficit |  | Pt referred for driving assessment |  |
| cognitive deficit |  |  | Independence on Disability Measures |
| Visual deficit |  | Return to work | Modified Rankin Scale* |
| perceptual deficit | | Patient asked if they want to RTW | FIM Cognition\$ |
| hydration problems |  | Did the patient want to return to work? | FIM Motor^ |
| nutrition problems |  | Informed of services to assist with RTW if wanting to RTW |  |
| Arm deficit |  | Was patient informed of services to assist with return to work | Change in FIM |
| at risk of malnutrition |  |  | FIM Motor^ |
| | | Communication and Peer Support on discharge | FIM Cognition\$ |
| Independence on Disability Measures |  | Offered info about sexuality after stroke |  |
| Modified Rankin Scale* |  | Patient provided info about peer support | Length of Stay |
| FIM Cognition\$ | | Carer provided with info re peer support | |
| FIM Motor^ |  | Discharge Summary provided to GP |  |
|  |  | Hospital contact for discharge programs given to patient |  |
|  |  | Hospital contact for discharge programs given to family |  |
|  |  | In hospital contact for discharge programs provided to patient OR family |  |
|  |  | Patient made aware of the availability of generic self-management programs before discharge from hospital |  |
| **bold font= p<0.05; † if impairment present ; \* defined as modified Rankin Scale score 0-2; \$ defined as Functional Independence Measure Cognitive score 30-35; ^ defined as Functional Independence Measure Motor score 78-91;** | | | |

